# SARS-CoV-2 Serology Status Detected by Commercialized Platforms Distinguishes Previous Infection and Vaccination Adaptive Immune Responses

**DOI:** 10.1101/2021.03.10.21253299

**Authors:** Raymond T. Suhandynata, Nicholas J. Bevins, Jenny T. Tran, Deli Huang, Melissa A. Hoffman, Kyle Lund, Michael J. Kelner, Ronald W. McLawhon, Steven L. Gonias, David Nemazee, Robert L. Fitzgerald

**Affiliations:** Department of Pathology UC San Diego Health, San Diego CA; Department of Immunology and Microbiology, The Scripps Research Institution, San Diego, CA

**Author notes:** Address Correspondence to: Robert Fitzgerald.

**Keywords:** Vaccines, Neutralizing Antibodies, COVID-19, Serology, SARS-CoV-2, Commercial Assays

## Abstract

**Background:** The severe acute respiratory syndrome coronavirus-2 (SARS-CoV-2) has infected over 110 million individuals and led to 2.5 million deaths worldwide. As more individuals are vaccinated, the clinical performance and utility of SARS-CoV-2 serology platforms needs to be evaluated.

**Methods:** The ability of four commercial SARS-CoV-2 serology platforms to detect previous infection or vaccination were evaluated using a cohort of 53 SARS-CoV-2 PCR-positive patients, 89 SARS-CoV-2-vaccinated healthcare workers (Pfizer or Moderna), and 127 SARS-CoV-2 negative patients. Serology results were compared to a cell based SARS-CoV-2 pseudovirus (PSV) neutralizing antibodies assay.

**Results:** The Roche S-(spike) antibody and Diazyme neutralizing antibodies (NAbs) assays detected adaptive immune response in 100.0% and 90.1% of vaccinated individuals who received two-doses of vaccine (initial and booster), respectively. The Roche N-(nucleocapsid) antibody assay and Diazyme IgG assay did not detect adaptive immune response in vaccinated individuals. The Diazyme Nabs assay correlated with the PSV SARS-CoV-2 ID50 neutralization titers (R^2^= 0.70), while correlation of the Roche S-antibody assay was weaker (R^2^= 0.39). Median PSV SARS-CoV-2 ID50 titers more than doubled in vaccinated individuals who received two-doses of the Moderna vaccine (ID50: 597) compared to individuals that received a single dose (ID50: 284).

**Conclusions:** The Roche S-antibody and Diazyme NAbs assays robustly detected adaptive immune responses in SARS-CoV-2 vaccinated individuals and SARS-CoV-2 infected individuals. The Diazyme NAbs assay strongly correlates with the PSV SARS-CoV-2 NAbs in vaccinated individuals. Understanding the reactivity of commercially available serology platforms is important when distinguishing vaccination response versus natural infection.

**Summary:** The Roche S (spike protein)-antibody and Diazyme neutralizing-antibodies (NAbs) assays were evaluated for their clinical utility in the detection of SARS-CoV-2 related adaptive immune responses by testing SARS-CoV-2 PCR-confirmed patients, SARS-CoV-2-vaccinated individuals, and SARS-CoV-2-negative individuals. Commercial serology results were compared to results generated using a cell-based SARS-CoV-2 pseudovirus (PSV) NAbs assay and previously validated SARS-CoV-2 commercial serology assays (Roche N (nucleocapsid protein) antibody and Diazyme IgG). We demonstrate that the Roche S-antibody and Diazyme NAbs assays detected adaptive immune response in SARS-CoV-2 vaccinated individuals and the presence of SARS-CoV-2 PSV NAbs. The Roche S-antibody assay had an observed positive percent agreement (PPA) of 100% for individuals who received two doses of the Pfizer or Moderna vaccine. By contrast, the Roche N assay and Diazyme IgG assay did not detect vaccine adaptive immune responses. Our findings also indicate that the Diazyme NAbs assay correlates strongly with the levels of SARS-CoV-2 ID50 neutralization titers using the PSV Nab assay in vaccinated individuals.

## Introduction

As of Feb 13^th^, 2021, the severe acute respiratory syndrome coronavirus 2 (SARS-CoV-2) has resulted in 110 million documented infections and nearly 2.5 million deaths worldwide (1). Recently, the United States Food and Drug Administration (USFDA) and European Union (EU) authorized use of SARS-CoV2 vaccines, including BNT162b2 (Pfizer-BioNTech) and mRNA-1273 (Moderna) (2,3). The Moderna and Pfizer vaccines utilize lipid nanoparticles that encapsulate RNA encoding the pre-fusion stabilized spike protein (S-protein) from SARS-CoV-2 (4,5). Previous *in vitro* studies have demonstrated that antibody-binding to specific S-protein epitopes disrupts viral entry and subsequent infection (6). Development of SARS-CoV-2 vaccines required amino acid substitutions in the recombinant prefusion conformation of S-protein to stabilize the structure allowing for better recognition by the humoral immune system (4,5,7). It was thus unclear whether commercial serology assays designed to recognize S-protein would detect antibodies generated in vaccinated patients.

Most of the original serology tests for SARS-CoV-2 were designed to detect patients who were previously exposed to the virus (8,9). Many of these assays target antibodies produced against the SARS-CoV-2 nucleocapsid (N) protein, with several others target a combination of the N and S proteins (10,11). Serology that exclusively detect N-protein antibodies would most likely not detect individuals vaccinated against the S-protein. The performance of serology tests that detect S-protein antibodies or a mixture of S and N protein antibodies in response to vaccination remains incompletely understood. Clinical interpretation of commercial serology results requires an understanding of the performance of these assays in both vaccinated individuals and those previously infected with SARS-CoV-2.

The relationship between SARS-CoV-2 serology results and protective immunity against SARS-CoV-2 has been a topic of great debate (9). Some critics argue that SARS-CoV-2 serology testing may create a false sense of protection amongst patients testing positive (12,13), while others argue that serology testing is an important epidemiological and clinical tool when used appropriately (14). Several studies have correlated commercial SARS-CoV-2 serology results to cell-based SARS-CoV-2 neutralizing antibody (Nab) assays (15–19). Unfortunately, these cell-based assays are generally unsuitable for use in a high-throughput clinical laboratory. However, a recently released proxy assay for SARS-CoV-2 Nabs (Diazyme) has been developed which measures the interaction between the S-protein and the human angiotensin converting enzyme 2 (ACE2) receptor; which is the virus’s mode of infection into host cells (20,21). This high-throughput commercialized assay may provide a method to determine whether patients have NAbs against SARS-CoV-2.

Herein, we evaluate the ability of the Roche Elecsys Anti-SARS-CoV-2 S-antibody test, the Roche Elecsys Anti-SARS-CoV-2 N-antibody test, the Diazyme Anti-SARS-CoV-2 IgG assay, and the Diazyme NAbs assay to detect and discriminate previously infected, vaccinated, and non-infected individuals. We also compared these commercial serology assays to a previously validated (16) cell-based SARS-CoV-2 pseudovirus (PSV) assay to determine if serology status predicts the level of NAbs produced by vaccinated individuals.

## Materials and Methods

### Study Design and Patient Cohort

275 serum and plasma specimens from 269 individuals were collected in BD Vacutainer collection tubes (K-EDTA, lithium-heparin plasma separator tubes, and/or serum separator tubes) (**Supplementary Fig. 1**). This included serum or plasma samples from 53 patients (53 specimens) who tested positive for SARS-CoV-2 by a PCR-based assay, 89 vaccinated healthcare workers who received the Moderna (61 individuals, 66 specimens) or Pfizer (28 individuals, 29 specimens) vaccine, or 116 control patient specimens (116 patients) that were collected in 2018 and had been stored at −20°C, and 11 patients (11 specimens) who tested positive for respiratory infections other than SARS-CoV-2 using a respiratory panel nucleic acid (RPNA) detection test. Specimens from PCR-confirmed Covid-19 patients were collected prior to August 1^st^, 2020 to eliminate the possibility that they had been vaccinated. The PCR confirmed positive samples (N=53) were separated into two time-frames relative to a positive SARS-CoV-2 PCR confirmed result: < 15-days post diagnosis by PCR (n = 25) and ≥ 15-days post diagnosis by PCR (n=28). All of the PCR confirmed positive samples came from COVID-19 patients who had sufficient symptoms to require hospitalization. The vaccinated healthcare worker group was a convenience cohort who all were subject to weekly PCR testing for at least one month prior to being vaccinated. All patient specimens were collected under UCSD IRB protocol 181656.

### Confirmation of SARS-CoV-2 Positive Patients

All 53 SARS-CoV-2 patients were positively confirmed for COVID-19 by an emergency use authorized (EUA) nucleic acid test validated in our laboratory. The platforms used included the Abbott ID NOW, Abbott m2000, Abbott Alinity, Abbott RealTime, Hologic Panther, ThermoFisher TaqPath, GenMark ePlex, and the Roche Cobas 6800/8800. All patients that tested positive using any of these platforms were considered SARS-CoV-2 PCR-confirmed patients and for discussion simplicity are referred to PCR positive.

### Commercial Serology and Neutralization Assays

Serology assays were performed on the Roche Cobas 8000 e801/e601 and included the Elecsys Anti-SARS-CoV-2 N-antibody test and the Elecsys Anti-SARS-CoV-2 S-antibody test. The Roche N-antibody assay reports a cutoff index (COI; signal of sample/cutoff); values ≥ 1.00 COI are considered reactive. The Roche S-antibody assay reports results in absorbance units per mL (AU/mL); values ≥ 0.8 AU/mL are considered reactive. A 1:10 dilution was performed in accordance with the manufacturer’s package insert for Roche S-antibody assay values >250 AU/mL which extends the analytical measurement range (AMR) of the assay to 2500 AU/mL. Additional serology testing was performed using the Diazyme DZ-Lite 3000 plus clinical analyzer (Diazyme DZ-LITE 2019-nCoV IgG and Diazyme SARS-CoV-2 NAbs) in accordance with the manufacturer’s instructions. The Diazyme IgG platform reports results in absorbance units per mL (AU/mL); values ≥ 1.00 AU/mL are considered reactive. The Diazyme NAbs assay reports results in absorbance units per mL (AU/mL); a manufacturer’s suggested cutoff is not provided with the package insert and was set at three standard deviations above the mean measured value of the 127 SARS-CoV-2 negative specimens (0.488 AU/mL).

### GenMark ePlex Respiratory Pathogen Nucleic Acid Test

To identify patient specimens containing other PCR confirmed microbes, a respiratory pathogen nucleic acid (RPNA) test was performed on the GenMark ePlex (GenMark Diagnostics Inc., Carlsbad, CA, USA). This panel detects Adenovirus (A-F), Coronavirus (229E, HKU1, NL63, OC42), Human Metapneumovirus, Human Rhinovirus/Enterovirus, Influenza A, B and C, Influenza 2009 H1N1, Parainfluenza (1-4), Respiratory Syncytial Virus (A and B), Chlamydia pneumoniae and Mycoplasma pneumoniae.

### Precision Studies

Precision was calculated across 5 days by running 5 batches of 5 replicates of patient pooled plasma (Roche S-antibody assay) or quality control material (Diazyme NAbs assay) (n=25). The positive patient pool was made by combining SARS-CoV-2 PCR positive patient samples and diluting the pool with SARS-CoV-2 negative patient serum to fall within the AMR of the assay. Negative patient pools were created by combining patient specimens (Li-heparin or K-EDTA) collected from control patients in 2018 that had been stored at −20° C.

### Dilutional linearity

Dilutional linearity for the Roche S-antibody and Diazyme NAbs assays was assessed in triplicate by mixing a positive patient pool with a negative patient pool in 10% increments prior to analysis. Patient pools for each assay were unique to account for differences in assay sensitivity.

### SARS-CoV-2 Pseudovirus Neutralization Assay

SARS-CoV-2 neutralization assays were performed as previously described using a pseudovirus (PSV) (16,22). In brief, the assay uses single cycle infectious viral particles containing firefly luciferase. The amount of luminescence in HeLa cells that stably expressed the cell surface receptor angiotensin converting enzyme 2 were measured after viral infection. Titers of 50% inhibitory dilution (ID50) were determined. For this study, the cutoff for a positive PSV neutralization result was raised to 100 (previously 50) to adjust for background changes in the assay, thus ID50 titers of greater than or equal to 100 were considered positive for PSV NAbs against SARS-CoV-2.

### Statistical Analyses

Data were analyzed using R in Rstudio. Linear regression analysis was performed in excel. Box plots were generated in Rstudio. Median values and inter-quartile ranges (IQR) are indicated for each box-plot. The upper whisker extends from the hinge to the largest value no further than 1.5 times the IQR from the hinge (where IQR is the inter-quartile range, or distance between the first and third quartiles). The lower whisker extends from the hinge to the smallest value at most 1.5 times the IQR of the hinge. Data beyond the end of the whiskers are called “outlying” points and are plotted individually. p-values were generated using a Wilcoxon test (p-value < 0.05 was considered significant). Precision (%CV) was calculated by analysis of variance (ANOVA) and total precision (%CV) was calculated by the sum of squares. PPA was defined as the number of positive test results divided by the sum of positives and negatives. NPA was defined as the number of negative test results divided by the sum of negatives and positives as has been described previously (15).

## Results

### Analytical Performance of Serology Tests

Within-run, between-run, and total precision (%CV) of the Roche S-antibody and Diazyme NAbs assays are presented in **Supplementary Table 1**. Within run precision ranged from 1.4 - 2.7% for the Roche S-antibody assay and from 3.6 - 7.4% for the Diazyme NAbs assay. Between-run precision ranged from 2.4 - 3.6% for the Roche S-antibody assay and from 1.6 - 1.9% for the Diazyme NAbs assay. Total precision ranged from 2.8 - 4.5% for the Roche assay and from 4.0 - 7.7% for the Diazyme NAbs assay. Precision characteristics of the Roche N-antibody and Diazyme IgG assays have been previously published (23,24).

Linear regression analysis was performed to illustrate the relationship between mean observed and expected values following sample dilution for the Roche S-antibody and Diazyme NAbs assays (**Supplementary Fig. 2**). Both the Roche S-antibody assay (R^2^= 0.996, y= 1.01x - 7.00) and the Diazyme NAbs assay (R^2^= 0.950, y= 1.08x + 0.98) diluted in a linear fashion (**Supplementary Fig. 2A and 2B**). Dilutional performance of the Roche N-antibody and Diazyme IgG assays have been previously reported (23,24).

Cross-reactivity of the Roche S-antibody and Diazyme NAbs assays was evaluated using specimens from 11 patients infected with other types of respiratory pathogens, including 7 with non-COVID-19 coronavirus. No cross-reactivity was observed for the Roche or Diazyme platforms as all 11 specimens were negative (**Supplementary Table 2**).

### Detection of SARS-CoV-2 Adaptive Immune Response in Infected Patients

The positive percent agreement (PPA) between commercial assays and a positive SARS-CoV-2 PCR result was performed by dividing PCR positive patients into two different time-frames relative to the time of diagnosis by a PCR test (**Table 1**). The < 15 day group included 25 PCR positive patients and the ≥ 15 day group included 28 PCR positive patients. The PPA for the < 15 day group was less than 100% across all platforms: Roche S-antibody (88.0%), Roche N-antibody (84.0%), Diazyme IgG (84.0%), and Diazyme NAbs (80.0%). By contrast, the PPA observed in the ≥ 15 day group was 100% for all four assays, illustrating the time-dependent performance of serology and Nab assays following infection. The negative percent agreement (NPA) for the serology assays in the PCR-negative cohort collected in 2018 was 100% for the Roche S-antibody and Diazyme IgG assays, 99.2% for the Roche N-antibody assay, and 98.4% for the Diazyme NAbs assay (**Table 1)**.

**Table 1:** PPA and NPA of Commercial Serology Platforms With SARS-CoV-2 PCR Positive Patients.

| **Positive Percent Agreement (PPA)** |  |  |  |  |
| --- | --- | --- | --- | --- |
| **SARS-CoV-2 PCR ( < 15 Days Post PCR Positive) n=25** |  |  |  |  |
| Metric | Roche S | Roche N | Diazyme IgG | Diazyme Nabs |
| PCR Positive | 25 | 25 | 25 | 25 |
| Positive on Assay | 22 | 21 | 21 | 20 |
| **PPA** | **88.0%** | **84.0%** | **84.0%** | **80.0%** |
| **Positive Percent Agreement (PPA)** |  |  |  |  |
| **SARS-CoV-2 PCR ( ≥ 15 Days Post PCR Positive) n=28** |  |  |  |  |
| Metric | Roche S | Roche N | Diazyme IgG | Diazyme Nabs |
| PCR Positive | 28 | 28 | 28 | 28 |
| Positive on Assay | 28 | 28 | 28 | 28 |
| **PPA** | **100.0%** | **100.0%** | **100.0%** | **100.0%** |
| **Negative Percent Agreement (NPA)** |  |  |  |  |
| **2018 Samples (n=116) and Other Respiratory Pathogens (n=11)** |  |  |  |  |
| Metric | Roche S | Roche N | Diazyme IgG | Diazyme Nabs |
| COVID-19 Negative | 127 | 127 | 127 | 127 |
| Positive on Assay | 0 | 1 | 0 | 2 |
| **NPA** | **100.0%** | **99.2%** | **100.0%** | **98.4%** |

The distribution of observed values on each commercial serology platform for SARS-CoV-2 PCR-positive patients is shown in **Supplementary Figure 3**. The observed values for patients tested < 15 days post-PCR positive were significantly lower compared to patients tested ≥ 15 days post-PCR positive (p-value ≤ 0.05).

### Detection of Adaptive Immune Response in Vaccinated Individuals

To evaluate whether the commercial serology assays detect SARS-CoV-2 vaccinated individuals (Pfizer or Moderna), the PPA between vaccinated individuals and a positive result on each commercial assay was determined (**Table 2**). Vaccinated individuals were divided into two groups based on whether an individual had or had not received their booster shot (Pre-booster vs Post-booster). The median number of days post-initial injection for pre-booster individuals was 22.5 days with an interquartile range (IQR) of 16.25 – 28.75 days. The median number of days post-initial injection for post-booster individuals was 37 days with an IQR of 34 – 38 days. The Roche S-antibody assay had an observed PPA of 87.5% for pre-booster individuals and an observed PPA of 100% for post-booster individuals. The Diazyme NAbs assay was less sensitive to detecting adaptive immune response related to vaccination status and had an observed PPA of 66.6% for pre-booster individuals and an observed PPA of 94.4% for post-booster individuals. The Roche N-antibody assay and Diazyme IgG assay did not react with vaccinated individuals. All vaccinated individuals depicted in Table 2 that were reactive on the Roche N-antibody and Diazyme IgG assays were previously infected with SARS-CoV-2 (PCR confirmed).

**Table 2:** PPA of Commercial Serology Platforms with SARS-CoV-2 Vaccinated Individuals.

| **Vaccine Positive Percent Agreement (PPA)**<br>**Specimens From Vaccinated Individuals (Pre-Booster)**<br>**Median Days Post-Initial Injection: 22.5 days**<br>**IQR: 16.25 -28.75 days** |  |  |  |  |
| --- | --- | --- | --- | --- |
| Metric | Roche S | Roche N | Diazyme IgG | Diazyme Nabs |
| Vaccinated | 24 | 24 | 24 | 24 |
| Positive on Assay | 21 | 1 | 1 | 16 |
| **PPA** | **87.5%** | **3.8%** | **3.8%** | **66.6%** |
| **Specimens From Vaccinated Individuals (Post-Booster)**<br>**Median Days Post-Initial Injection: 37 days**<br>**IQR: 34 -38 days** |  |  |  |  |
| Metric | Roche S | Roche N | Diazyme IgG | Diazyme Nabs |
| Vaccinated | 71 | 71 | 71 | 71 |
| Positive on Assay | 71 | 2 | 1 | 67 |
| **PPA** | **100.0%** | **2.8%** | **1.4%** | **94.4%** |

The median value observed for vaccinated individuals on the Diazyme IgG assay was 0.039 AU/mL (IQR: 0.024 – 0.069) for pre-booster individuals and 0.044 AU/mL (IQR: 0.028 – 0.060) for post-booster individuals (**Fig. 1A**). The median value observed on the Roche N-antibody assay was 0.077 COI (IQR: 0.076 – 0.079) for pre-booster individuals and 0.077 COI (IQR: 0.075 – 0.081) for post-booster individuals (**Fig. 1B**). The median value observed for vaccinated individuals on the Diazyme NAbs assay was 0.738 AU/mL (IQR: 0.478 – 1.769) for pre-booster individuals and 3.109 AU/mL (IQR: 1.659 – 10.43) for post-booster individuals (**Fig. 1C**). The median value observed for vaccinated individuals on the Roche S-antibody assay was 62.3 AU/mL (IQR: 38.07 – 210.88) for pre-booster individuals and >2500 AU/mL (IQR: 1009 – >2500) for post-booster individuals (**Fig. 1D**). Median values observed for the Diazyme IgG and Roche N-antibody assays were well below the cutoff for a positive test result with no significant difference between pre-booster and post-booster individuals. In contrast, for both pre-booster and post-booster individuals, the median values observed for the Diazyme NAbs and Roche S-antibody assays were above the cutoff for a positive result and were significantly different (pre to post booster p-value < 0.05).

**Figure 1:**
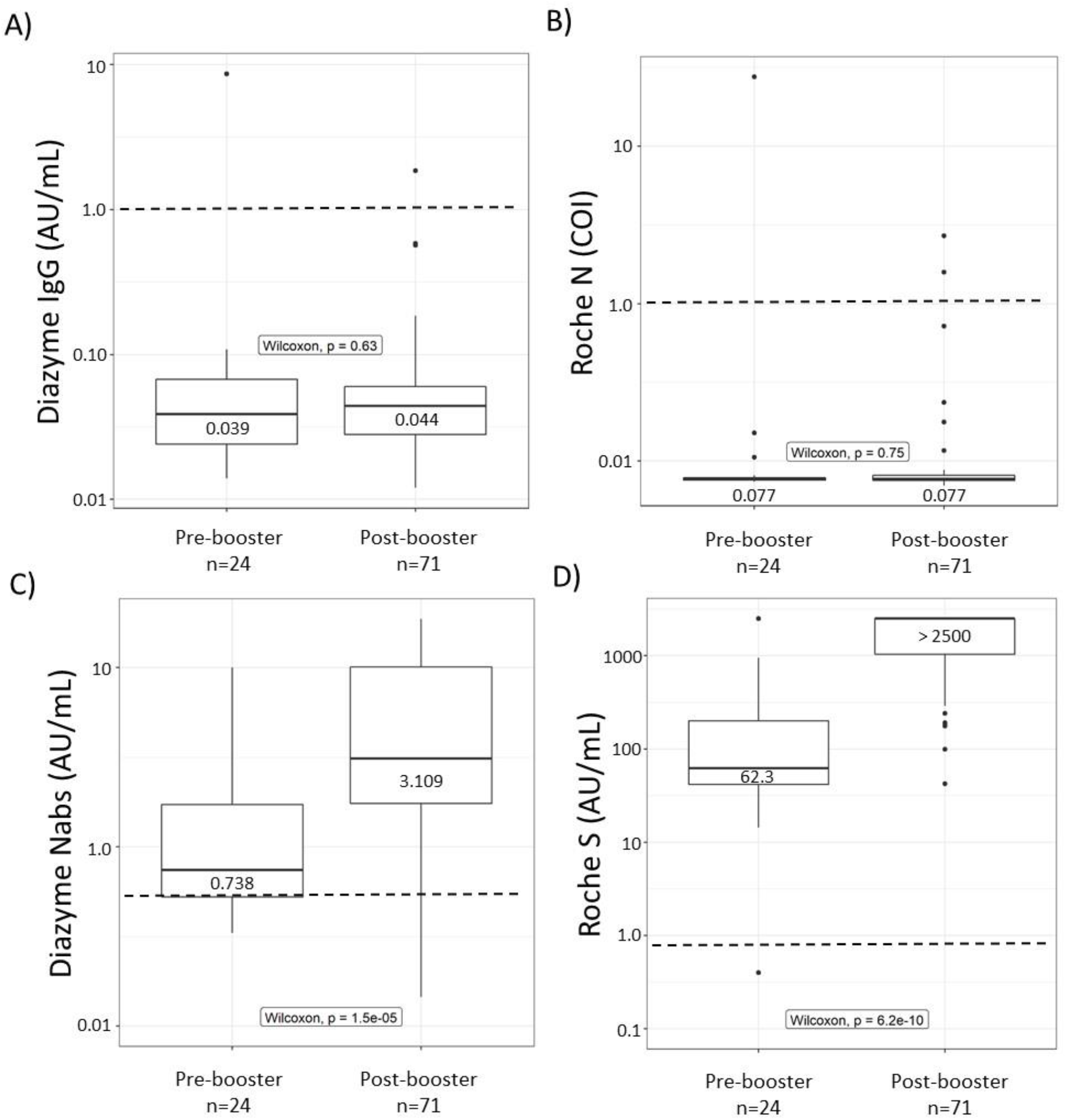
Commercialized Platforms Detect Vaccinated Individuals. **A-D**) Box-plots illustrating the distribution of values observed on the Diazyme IgG, Roche N-antibody, Diazyme Nabs, and Roche S-antibody assays. Vaccinated individuals are stratified based on whether or not they had received a booster shot (Pre-booster vs Post-booster). Observed values are plotted on the Y-axis on a Log_10_ scale. The dashed line indicates the cutoff for each platform with values above representing positive results and values below indicating negative results. Median values and interquartile ranges are indicated for each box-plot. Whiskers are up to 1.5 times the interquartile range.

### Correlation of Commercial Serology Results to SARS-CoV-2 ID50 Neutralization Titers

Utilizing a clinically validated cell-based PSV assay (15), we measured Nab titers (ID50) against SARS-CoV-2 in vaccinated individuals. The PPA between vaccinated individuals and the SARS-CoV-2 PSV assay was 87.5% for pre-booster individuals and 98.6% for post-booster individuals (**Table 3**). Furthermore, the median ID50 titer observed in pre-booster individuals was 285 (IQR: 141 – 485), while the median ID50 titer more than doubled to 597 (IQR: 359 – 1032) for post-booster individuals. Stratifying post-booster individuals based on vaccine manufacturer reveals that Moderna vaccinated individuals’ median ID50 titers were not significantly different than Pfizer vaccinated individuals; 597 for Moderna and 599 for Pfizer (**Fig 2A and 2B**). There was a clear difference between pre and post-booster ID50 titers for Moderna vaccinated individuals (p-value < 0.05) (**Fig 2A**). Significant difference was not observed between ID50 titers produced in pre-booster vs post-booster Pfizer vaccinated individuals (**Fig 2B**), where a robust response was seen after the first dose, although the number of subjects in this group was small (N=5). A strong correlation to SARS-CoV-2 ID50 neutralization titers was observed with the Diazyme NAbs assay (R^2^= 0.70), while a weaker correlation was observed for the Roche S-antibody assay (R^2^= 0.39) (**Fig. 2C and 2D**).

**Table 3:**
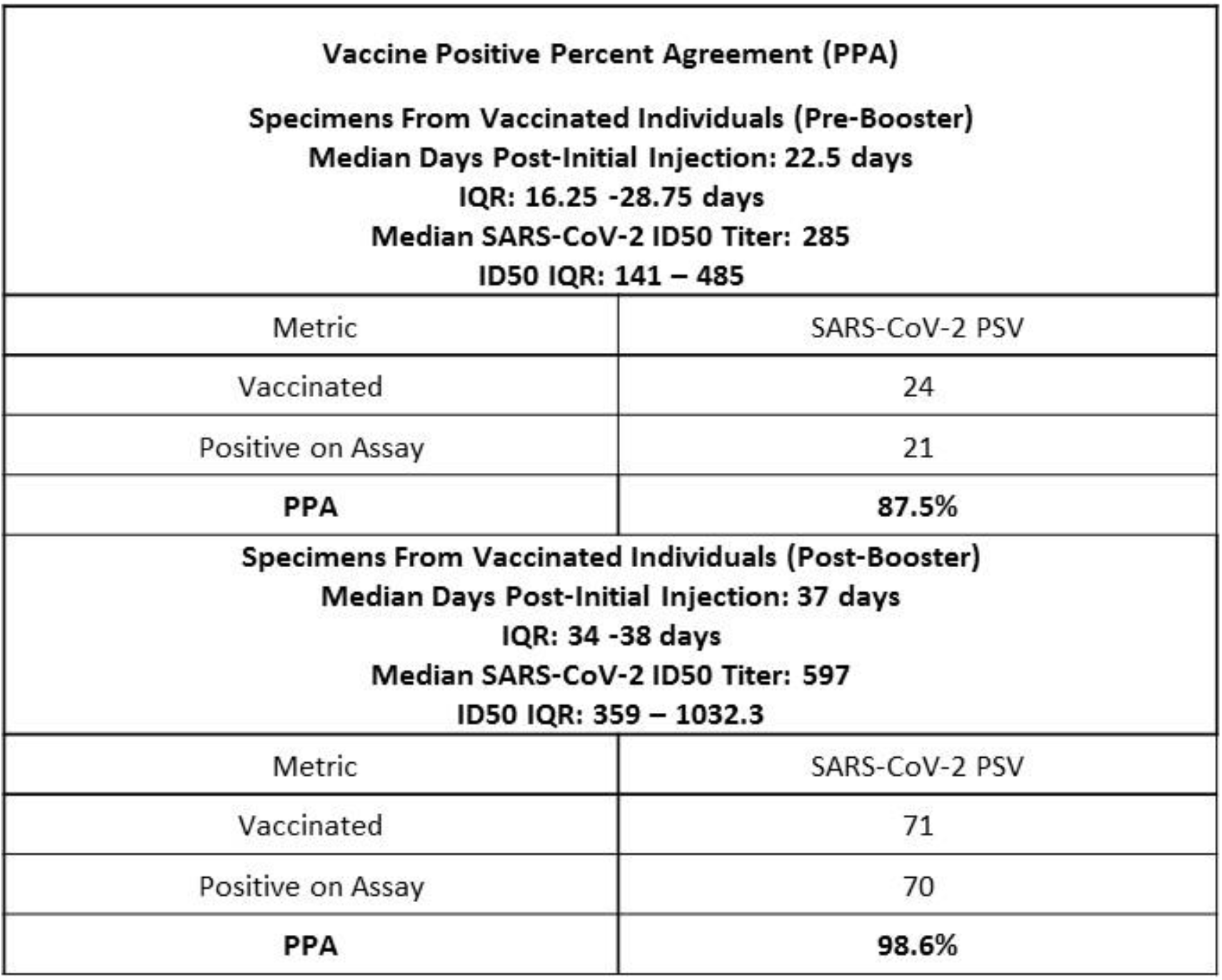
PPA of the SARS-CoV-2 PSV Assay with Vaccinated Individuals.

|  |  |
| --- | --- |
| **Vaccine Positive Percent Agreement (PPA)**<br>**Specimens From Vaccinated Individuals (Pre-Booster)**<br>**Median Days Post-Initial Injection: 22.5 days**<br>**IQR: 16.25 -28.75 days**<br>**Median SARS-CoV-2 ID50 Titer: 285**<br>**ID50 IQR: 141 – 485** |  |
| Metric | SARS-CoV-2 PSV |
| Vaccinated | 24 |
| Positive on Assay | 21 |
| **PPA** | **87.5%** |
| **Specimens From Vaccinated Individuals (Post-Booster)**<br>**Median Days Post-Initial Injection: 37 days**<br>**IQR: 34 -38 days**<br>**Median SARS-CoV-2 ID50 Titer: 597**<br>**ID50 IQR: 359 – 1032.3** |  |
| Metric | SARS-CoV-2 PSV |
| Vaccinated | 71 |
| Positive on Assay | 70 |
| **PPA** | **98.6%** |

**Figure 2:**
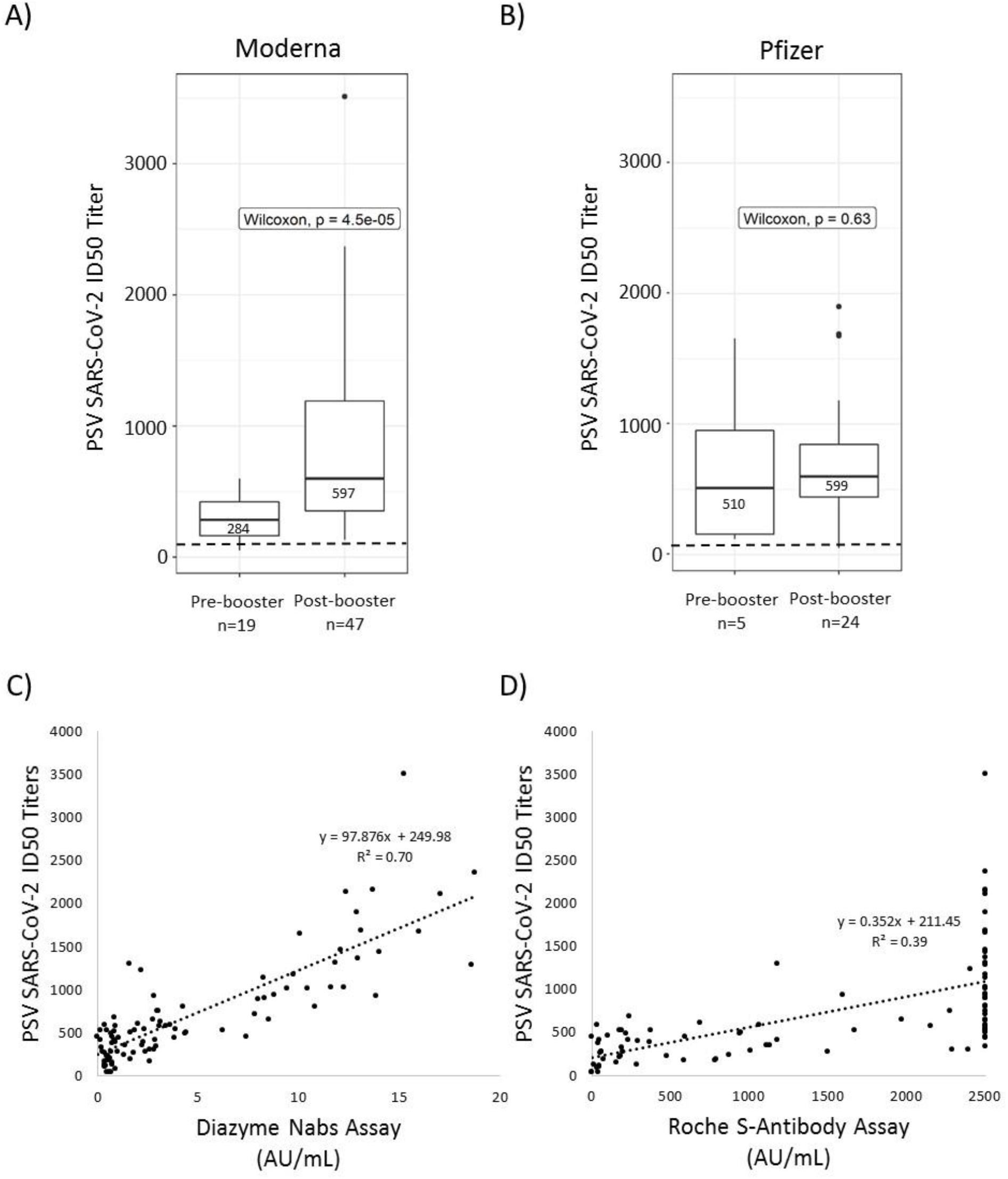
Nabs In Vaccinated Individuals and Correlation To Commercial Platforms. **A)** Box-plot describing the distribution of SARS-CoV-2 ID50 titers (Y-axis) across vaccinated individuals receiving the Moderna vaccine. **B**) Box-plot describing the distribution of SARS-CoV-2 ID50 titers (Y-axis) across vaccinated individuals receiving the Pfizer vaccine. The positive cutoff for Nabs on the PSV assay is indicated by a dashed line. Median values and interquartile ranges are indicated for each box-plot. Whiskers are up to 1.5 times the interquartile range. **C, D**) Linear regression analysis of SARS-CoV-2 ID50 Titers (Y-axis) observed in vaccinated individuals compared to values observed on the Diazyme Nabs or Roche S-antibody assays (X-axis). Linear regression analysis with corresponding R^2^ values and y=b + mx equations are indicated within each plot.

### Temporal Evaluation of SARS-CoV-2 NAbs and Serology in Vaccinated Individuals

The appearance of PSV SARS-CoV-2 NAbs is shown relative to an individual’s initial vaccination date (**Fig. 3A and 3B**). For pre-booster vaccinated individuals, ID50 titers ranged from 49 - 952 (**Fig. 3A**). As expected, the highest ID50 titers were observed in post-booster individuals (**Fig. 3B**), with PSV ID50 titers ranging from 49 – 3512. Three pre-booster individuals were negative for PSV SARS-CoV-2 NAbs. These individuals received the Moderna vaccine and were 7, 13, and 21 days post-initial injection. A single post-booster Pfizer vaccinated individual had PSV ID50 titers of <100 and was tested 24 days after their initial vaccination date and 3 days after receiving their booster. The earliest positive serology result for a vaccinated individual on either the Roche S-antibody or Diazyme NAbs assays was 12 days following initial injection of the vaccine (**Fig. 3C and 3D**).

**Figure 3:**
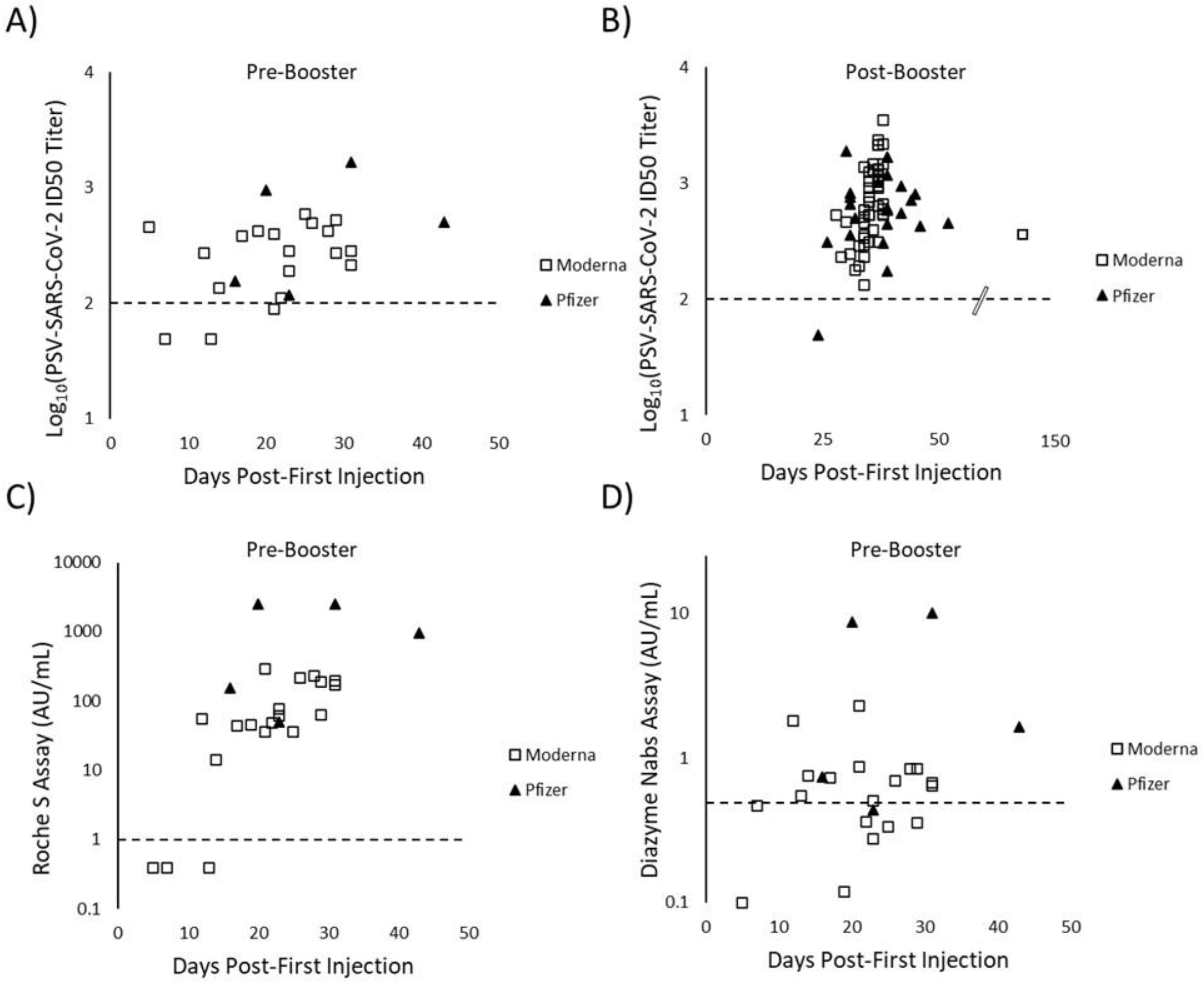
Kinetics of SARS-CoV-2 Nabs and Serology in Vaccinated Individuals. **A**) Log_10_(PSV SARS-CoV-2 ID50 neutralization titers) are shown (Y-axis) for pre-booster vaccinated individuals. **B**) Log_10_(PSV SARS-CoV-2 ID50 neutralization titers) are shown (Y-axis) for post-booster vaccinated individuals. The scale break-point on the X-axis is indicated for when the axis is no longer to scale. **C**) Values on the Roche S-antibody assay (Y-axis) for pre-booster vaccinated individuals. **D**) Values on the Diazyme Nabs assay (Y-axis) for pre-booster vaccinated individuals. For all graphs, individuals receiving the Moderna vaccine are indicated by open boxes while individuals receiving the Pfizer vaccine are indicated with solid triangles. The number of days following an individual’s initial vaccination date is plotted on the X-axis. The dashed X-axis indicates the positive cutoff for each assay.

## Discussion

The goal of the current study was determine the potential clinical utility of various SARS-CoV-2 antibody detection assays to identify and distinguish adaptive immune responses from prior SARS-CoV2 infection and vaccination. We compared four commercially available *in vitro* diagnostic serology assays and a cell-based assay, applicable mainly for research purposes. Each assay emerged as useful in specific contexts. The Roche S-antibody test and the Diazyme NAbs assay had appropriate precision for clinical use (**Supplementary Table 1)**. Both assays diluted in a linear fashion, with R^2^ values of greater than 0.95 (**Supplementary Fig. 2**). Previously we reported that the Abbott IgG and Roche N-antibody assays dilute in a non-linear fashion (23). Linear dilution is particularly critical when utilizing the Roche S-antibody assay because a large portion of the values observed on this platform were above the AMR (250 AU/mL) in both vaccinated individuals and SARS-CoV-2 infected patients.

The Roche S-antibody test and the Diazyme NAbs assay detect SARS-CoV-2 infected patients with similar sensitivity (PPA) and specificity (NPA) as previously validated SARS-CoV-2 serology assays (23–25); all four commercial serology platforms had PPAs of 100% for patients tested ≥ 15 days a post-PCR positive result. Since the Roche S-antibody and Roche N-antibody assays are performed on the same platform, clinical laboratories can utilize a single analyzer to implement the CDC’s recommended screen and confirm approach when testing low disease prevalence populations for past exposure to SARS-CoV-2 (26).

With the arrival of SARS-CoV-2 vaccines, a major concern is whether commercial SARS-CoV-2 serology assays can distinguish vaccinated individuals from those with natural infections. This is particularly important for patients with symptoms of “long haul” COVID-19 (27) and when diagnosing previously asymptomatic infections. The Roche S-antibody assay detected 100% of post-booster vaccinated individuals, making it an excellent choice for determining if vaccinated individuals have mounted an immune response **(Table 2)**. However, at the present time, antibody testing is not recommended by the CDC to confirm seroconversion following routine vaccinations (28). The Diazyme NAbs assay detected 94.4% of post-booster individuals, suggesting that it is either less sensitive than the Roche S-antibody assay or that some individuals produce antibodies to the S protein that are not neutralizing. The Roche N-antibody assay and Diazyme IgG assay did not detect vaccinated individuals, as all positive results from vaccinated individuals had a previous SARS-CoV-2 PCR confirmed infections. A combination of S-antibody and N-antibody assays can be used to differentiate naturally infected individuals from vaccinated individuals, as naturally infected individual are positive on both the S and N-antibody assays we evaluated. Furthermore, this strategy would likely also be useful for differentiating natural infection from vaccination with adenovirus based vaccines such as vaccines manufactured by AstraZeneca (29) and Johnson and Johnson (30); as these vaccines are also based on expression of recombinant pre-fusion stabilized S-protein. The distribution of values observed on the Diazyme NAbs and Roche S-antibody assays were significantly different between pre-booster and post-booster vaccinated individuals (p-value < 0.0001), which is the anticipated response.

The high PPAs observed between individuals receiving a vaccine and the SARS-CoV-2 PSV assay (87.5% for pre-booster individuals and 98.6% for post-booster individuals), indicates that 98.6 percent of vaccinated individuals developed NAbs against SARS-CoV-2 (**Table 3**). The median PSV SARS-CoV-2 ID50 titers were not significantly different between post-booster individuals that received the Moderna (597) or Pfizer (599) vaccines (**Fig 2 A and 2B**).

One limitation of our study is that the ID50 NAb titer that corresponds to immunity against SARS-CoV-2 infection is not known and may be different for NAbs targeting different epitopes (31). Previous studies of the relationship between serology and protection from non-SARS-CoV-2 coronaviridae have yielded varying results with high anti-viral IgG titers generally correlating with protection from severe clinical disease and reduced transmission (32). A commercially available assay that does not require a pseudoviral component and detects the presence and titer of NAbs will be important. A previous study indicates that the Roche N-antibody and Diazyme IgG assays had weak correlations with results from SARS-CoV-2 PSV Nab assay, with correlation coefficients of 0.27 and 0.40, respectively (16). The Diazyme NAbs assay strongly correlated with the level of SARS-CoV-2 ID50 PSV neutralization titers (R^2^=0.70) (**Fig. 2C)**, demonstrating the importance of functional assays and that not all antibodies made in response to vaccination have neutralizing activity.

As more individuals become vaccinated, it is important to understand the timeframe for when a vaccinated individual may be detected by these commercial assays. Our observations suggest that a healthcare provider should wait at least 2-weeks and preferably 3 weeks after an individual has received a SARS-CoV-2 vaccine before attempting to determine if they have produced antibodies, as the earliest observed seroconversion for individuals receiving the Moderna or Pfizer vaccines was 12 days and 16 days, respectively (**Fig. 3C and 3D**). These results correlate well with the kinetics of ID50 neutralization titers that vaccinated individuals generate in response to vaccination (**Fig. 3A and 3B**).

In summary, our study demonstrates that the Roche S-antibody and Diazyme NAbs assays detect responses to the Moderna and Pfizer SARS-CoV-2 vaccines as well as from natural infections. Both assays perform well compared to previously validated commercial SARS-CoV-2 serology platforms and correlate with the presence of NAbs against SARS-CoV-2, as determined by a cell-based PSV assay.

## Data Availability

All data is shown in the manuscript or in the supplemental files.

## Acknowledgement

We would like to acknowledge all the staff at the UC San Diego Health clinical laboratories for their help identifying specimens for assay validation and testing. We would also like to thank Amy Rockefeller and Ernestine Ferrer for valuable technical expertise. Waters Corporation and Roche Diagnostics are acknowledged for funding the clinical chemistry fellowships for R.T.S. and M.A.H respectively. D.H. and D.N. received support from NIH grants R37AI059714 and R01AI132317.

## Notes

### Competing Interest Statement

Diazyme provided evaluation kits for the proxy neutralizing antibody assay free of charge. They had no input on the design of the study, data analysis or writing of the manuscript. Data was not shared with Diazyme prior to submission of this manuscript.

### Author Declarations

All patient specimens were collected under UCSD IRB protocol 181656.

## References

1. Coronavirus Update (Live): 110,211,925 Cases and 2,434,029 Deaths from COVID-19 Virus Pandemic - Worldometer [Internet]. [cited 2021 Feb 17]. Available from: https://www.worldometers.info/coronavirus/

2. Commissioner O of the. COVID-19 Vaccines. FDA [Internet]. FDA; 2021 [cited 2021 Feb 17]; Available from: https://www.fda.gov/emergency-preparedness-and-response/coronavirus-disease-2019-covid-19/covid-19-vaccines

3. Safe COVID-19 vaccines for Europeans [Internet]. Eur. Comm. - Eur. Comm. [cited 2021 Feb 14]. Available from: https://ec.europa.eu/info/live-work-travel-eu/coronavirus-response/safe-covid-19-vaccines-europeans_en

4. Polack FP, Thomas SJ, Kitchin N, Absalon J, Gurtman A, Lockhart S, et al. Safety and Efficacy of the BNT162b2 mRNA Covid-19 Vaccine. N Engl J Med. 2020;383:2603–15.

5. Baden LR, El Sahly HM, Essink B, Kotloff K, Frey S, Novak R, et al. Efficacy and Safety of the mRNA-1273 SARS-CoV-2 Vaccine. N Engl J Med. 2021;384:403–16.

6. Brouwer PJM, Caniels TG, Aldon Y, Bangaru S, Torres JL, Okba NMA, et al. Potent neutralizing antibodies from COVID-19 patients define multiple targets of vulnerability. 2020;9.

7. Hsieh C-L, Goldsmith JA, Schaub JM, DiVenere AM, Kuo H-C, Javanmardi K, et al. Structure-based design of prefusion-stabilized SARS-CoV-2 spikes. Science. 2020;369:1501–5.

8. Vashist SK. In Vitro Diagnostic Assays for COVID-19: Recent Advances and Emerging Trends. Diagnostics. 2020;10:202.

9. Theel ES, Slev P, Wheeler S, Couturier MR, Wong SJ, Kadkhoda K. The Role of Antibody Testing for SARS-CoV-2: Is There One? J Clin Microbiol [Internet]. 2020 [cited 2021 Feb 17];58. Available from: https://www.ncbi.nlm.nih.gov/pmc/articles/PMC7383527/

10. Health C for D and R. EUA Authorized Serology Test Performance. FDA [Internet]. FDA; 2020 [cited 2020 Jul 27]; Available from: https://www.fda.gov/medical-devices/emergency-situations-medical-devices/eua-authorized-serology-test-performance

11. COVID-19 In Vitro Diagnostic Medical Devices | COVID-19 In Vitro Diagnostic Devices and Test Methods Database [Internet]. [cited 2021 Feb 17]. Available from: https://covid-19-diagnostics.jrc.ec.europa.eu/devices

12. Abbasi J. The Promise and Peril of Antibody Testing for COVID-19. JAMA. 2020;323:1881.

13. Torres R, Rinder HM. Double-Edged SpikeAre SARS-CoV-2 Serologic Tests Safe Right Now? Am J Clin Pathol. Oxford Academic; 2020;153:709–11.

14. Weinstein MC, Freedberg KA, Hyle EP, Paltiel AD. Waiting for Certainty on Covid-19 Antibody Tests — At What Cost? N Engl J Med. 2020;383:e37.

15. Jää skelä inen A, Kuivanen S, Kekä lä inen E, Ahava M, Loginov R, Kallio-Kokko H, et al. Performance of six SARS-CoV-2 immunoassays in comparison with microneutralisation. J Clin Virol. 2020;129:104512.

16. Suhandynata RT, Hoffman MA, Huang D, Tran JT, Kelner MJ, Reed SL, et al. Commercial Serology Assays Predict Neutralization Activity against SARS-CoV-2. Clin Chem. 2021;67:404–14.

17. Luchsinger LL, Ransegnola BP, Jin DK, Muecksch F, Weisblum Y, George PJ, et al. Serological Assays Estimate Highly Variable SARS-CoV-2 Neutralizing Antibody Activity in Recovered COVID-19 Patients. J Clin Microbiol. 2020;58:14.

18. Tang MS, Case JB, Franks CE, Chen RE, Anderson NW, Henderson JP, et al. Association between SARS-CoV-2 Neutralizing Antibodies and Commercial Serological Assays. Clin Chem. 2020;66:1538– 47.

19. Muecksch F, Wise H, Batchelor B, Squires M, Semple E, Richardson C, et al. Longitudinal Serological Analysis and Neutralizing Antibody Levels in Coronavirus Disease 2019 Convalescent Patients. J Infect Dis. 2021;223:389–98.

20. Hoffmann M, Kleine-Weber H, Schroeder S, Krüger N, Herrler T, Erichsen S, et al. SARS-CoV-2 Cell Entry Depends on ACE2 and TMPRSS2 and Is Blocked by a Clinically Proven Protease Inhibitor. Cell. 2020;181:271-280.e8.

21. Walls AC, Park Y-J, Tortorici MA, Wall A, McGuire AT, Veesler D. Structure, Function, and Antigenicity of the SARS-CoV-2 Spike Glycoprotein. Cell. 2020;181:281-292.e6.

22. Rogers TF, Zhao F, Huang D, Beutler N, Burns A, He W, et al. Isolation of potent SARS-CoV-2 neutralizing antibodies and protection from disease in a small animal model. Science. 2020;eabc7520.

23. Suhandynata RT, Hoffman MA, Kelner MJ, McLawhon RW, Reed SL, Fitzgerald RL. Multi-platform Comparison of SARS-CoV-2 Serology Assays for the Detection of COVID-19. J Appl Lab Med. 2020;jfaa139.

24. Suhandynata RT, Hoffman MA, Kelner MJ, McLawhon RW, Reed SL, Fitzgerald RL. Longitudinal Monitoring of SARS-CoV-2 IgM and IgG Seropositivity to Detect COVID-19. J Appl Lab Med. 2020;5:908–20.

25. Hubbard JA, Geno KA, Khan J, Szczepiorkowski ZM, de Gijsel D, Ovalle AA, et al. Comparison of Two Automated Immunoassays for the Detection of SARS-CoV-2 Nucleocapsid Antibodies. J Appl Lab Med. 2020;jfaa175.

26. CDC. Information for Laboratories about Coronavirus (COVID-19) [Internet]. Cent. Dis. Control Prev. 2020 [cited 2020 Jun 5]. Available from: https://www.cdc.gov/coronavirus/2019-ncov/lab/resources/antibody-tests-guidelines.html

27. Rubin R. As Their Numbers Grow, COVID-19 “Long Haulers” Stump Experts. JAMA. 2020;324:1381.

28. Interim Clinical Considerations for Use of mRNA COVID-19 Vaccines | CDC [Internet]. 2021 [cited 2021 Mar 3]. Available from: https://www.cdc.gov/vaccines/covid-19/info-by-product/clinical-considerations.html

29. ADIMITROVA EK. COVID-19 Vaccine AstraZeneca [Internet]. Eur. Med. Agency. 2021 [cited 2021 Mar 3]. Available from: https://www.ema.europa.eu/en/medicines/human/EPAR/covid-19-vaccine-astrazeneca

30. Janssen COVID-19 Emergency Use Authorization (EUA) Official Website [Internet]. [cited 2021 Mar 3]. Available from: https://www.janssencovid19vaccine.com/

31. Mor M, Werbner M, Alter J, Safra M, Chomsky E, Lee JC, et al. Multi-clonal SARS-CoV-2 neutralization by antibodies isolated from severe COVID-19 convalescent donors. Subbarao K, editor. PLOS Pathog. 2021;17:e1009165.

32. Huang AT, Garcia-Carreras B, Hitchings MDT, Yang B, Katzelnick LC, Rattigan SM, et al. A systematic review of antibody mediated immunity to coronaviruses: antibody kinetics, correlates of protection, and association of antibody responses with severity of disease. :47.

