## Supplemental Files for "SARS-CoV-2 Serology Status Detected by Commercialized Platforms Distinguishes Previous Infection and Vaccination Adaptive Immune Responses"

### Supplementary Table 1

**Supplementary Table 1: Precision Characteristics of the Roche S-antibody and Diazyme Nabs Assays**

| Sample (n=25) (Mean AU/mL, SD) | Roche S-antibody Assay | | |
|---|---|---|---|
| | **Between-run %CV** | **Within-run %CV** | **Total %CV** |
| Patient Pool Positive (1.74, 0.028) | 2.4 | 1.4 | 2.8 |
| Patient Pool Negative (0.55, 0.015) | 3.6 | 2.7 | 4.5 |
| **Sample (n=25) (Mean AU/mL, SD)** | **Diazyme Nabs** | | |
| | **Between-run %CV** | **Within-run %CV** | **Total %CV** |
| QC Level 2 (6.51, 0.445) | 1.9 | 7.4 | 7.7 |
| QC Level 1 (1.90, 0.064) | 1.6 | 3.6 | 4.0 |

### Supplementary Table 2

**Supplementary Table 2: Cross-reactivity of Commercial Assays to Patients with Other Respiratory Pathogens**

| Pathogen | Number of Samples Tested n=11 | Roche S-Antibody Assay | Roche N-Antibody Assay | Diazyme IgG Assay | Diazyme Nabs Assay |
|---|---|---|---|---|---|
| | | Number of Reactive | | | |
| Non COVID Coronavirus | 7 | 0 | 0 | 0 | 0 |
| Rhinovirus/ Enterovirus | 4 | 0 | 0 | 0 | 0 |

Supplementary Figure 1

A)

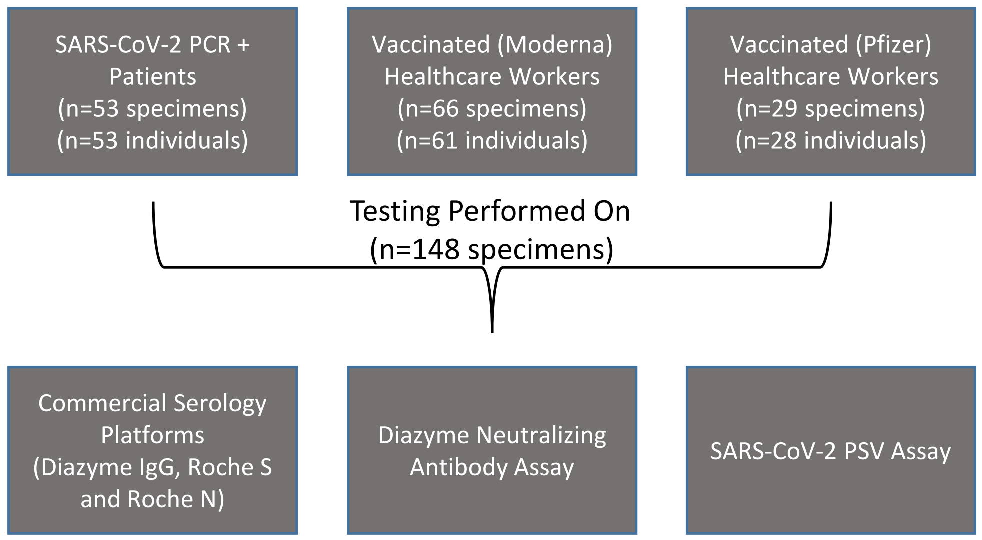


B)

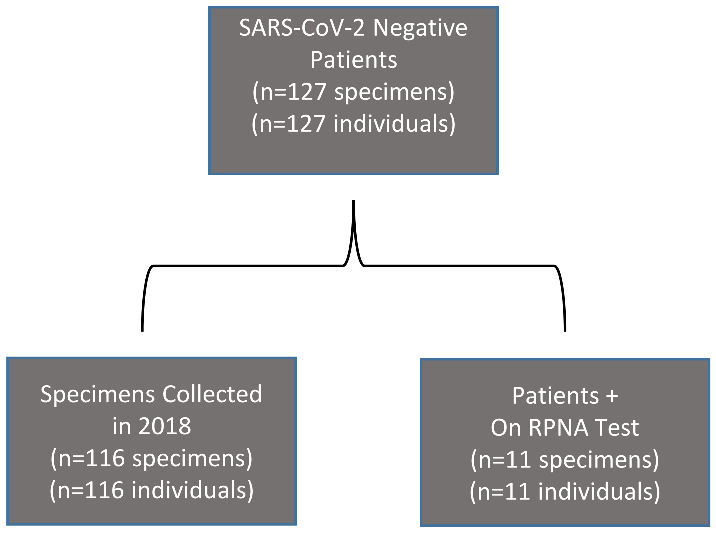


**Supplemantary Figure 1: Cohort Summary**
Flowcharts of the cohorts used for **A)** SARS-CoV-2 PCR positive and SARS-CoV-2 vaccinated healthcare workers. **B)** SARS-CoV-2 negative patients.

### Supplementary Figure 2

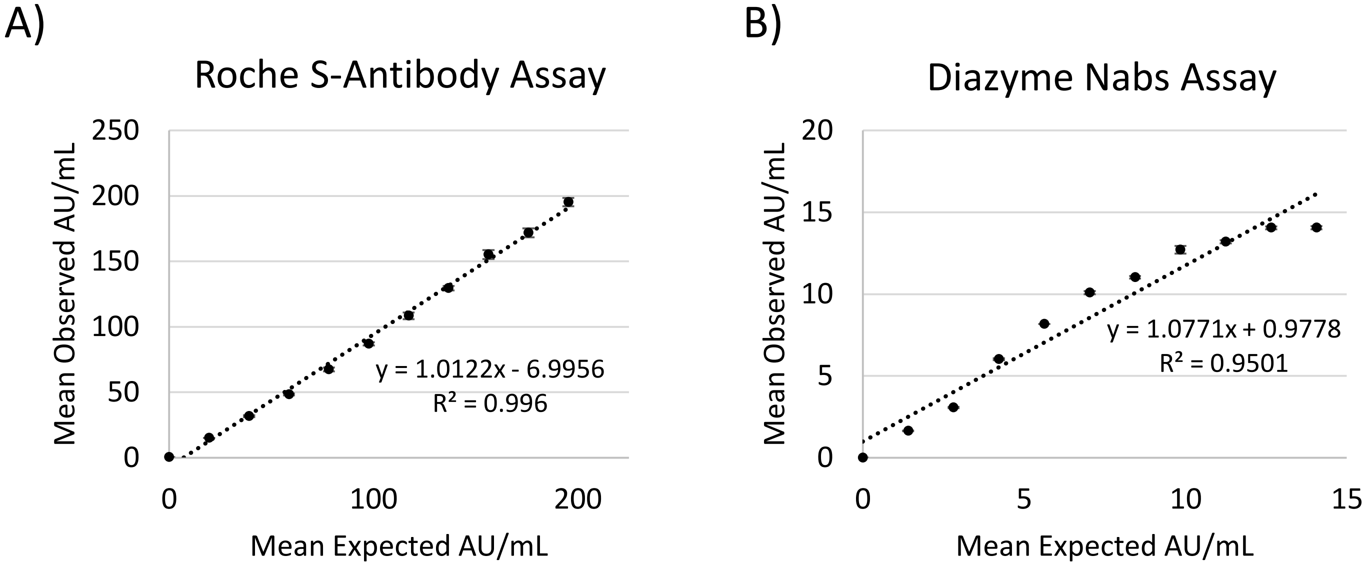


**Supplementary Figure 2: Dilution Performance of Roche S-antibody and Diazyme Nabs Assays**

**A)** Linear regression comparing observed mean AU/mL and expected AU/mL of a patient pool diluted in 10% increments on the Roche S-antibody assay. Analysis was performed in triplicate and error bars represent the standard error of the mean. **B)** Linear regression comparing the observed mean AU/mL and the expected mean AU/mL of a patient pool diluted in 10% increments on the Diazyme Nabs assay. Analysis was performed in triplicate and error bars represent the standard error of the mean. Linear regression analysis with corresponding $R^2$ values and y=b + mx equations are indicated within each plot

**Supplementary Figure 3: Commercialized Platforms Detect SARS-CoV-2 Infected Patients**

**A -D**) Box-plots illustrating the distribution of values observed on the Diazyme IgG, Roche N-antibody, Diazyme Nabs, and Roche S-antibody assays. SARS-CoV-2 PCR confirmed patients are stratified based on the number of days their samples were collected following a positive SARS-CoV-2 PCR result (< 15 days post-PCR + vs ≥ 15 days post-PCR +). Values observed are plotted on the Y-axis on a $Log_{10}$ scale and dashed red line indicates the cutoff for each platform with values above representing positive results and values below indicating negative results. Median values and interquartile ranges are indicated for each box-plot. Whiskers are up 1.5 times the interquartile range.

Supplementary Figure 3

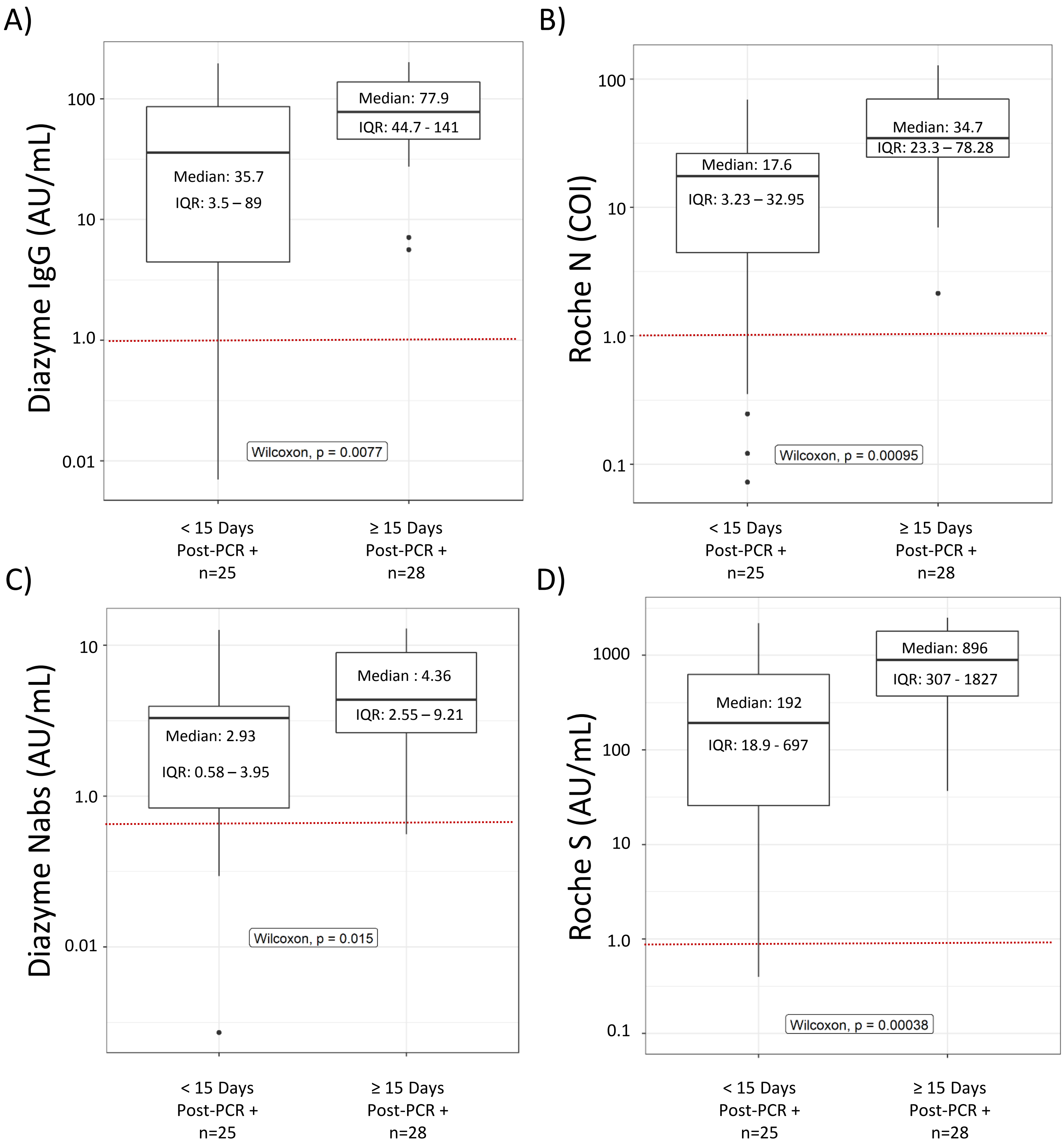
